# Long term stability of patients undergoing endovascular parent artery occlusion of their intracranial artery

**DOI:** 10.1101/2023.05.06.23289239

**Authors:** Satoshi Koizumi, Masaaki Shojima, Takahiro Ota, Shogo Dofuku, Satoru Miyawaki, Satoshi Kiyofuji, Keiichiro Maeda, Takashi Ochi, Akihiro Ito, Yukihiro Hidaka, Soichi Oya, Akira Saito, Gakushi Yoshikawa, Kei Yanai, Tomohiro Inoue, Sho Tsunoda, Katsumi Hoya, Nobuhito Saito

## Abstract

**Background:** Although endovascular parent artery occlusion (PAO) of the intracranial artery is a well-established treatment option, the long-term stability of cerebral blood flow remains a concern. This study aimed to evaluate the long-term clinical and radiological outcomes of patients who underwent PAO.

**Methods:** The patients who underwent endovascular PAO of their internal carotid or vertebral artery (VA) between April 2011 and March 2022 were included in this observational study. Information about patient characteristics, details of the endovascular treatment, and clinical and radiological follow-up were collected.

**Results:** The study included a total of 104 cases (average age 52.9±12.6 years old, male 73 (70.2%) cases, 95 (91.3%) VA PAO cases) from eight centers. Most cases were performed in an emergency condition, such as ruptured vertebral artery dissecting aneurysm (73 cases [70.2%]). PAO was successful in all cases. Early stroke (within 30 days) occurred in 33 (31.7%) cases (31 cases in VA PAO and two cases in internal carotid PAO) with ischemic stroke (29 cases) comprising the largest group. Clinical follow-up over 12 months was available in 78 cases. During an average follow-up period of 49.5 ± 24.3 months, one case in VA PAO experienced a stroke without functional deterioration. Imaging follow-up was performed in 73 cases. Recanalization of the occluded VA was observed in two cases. The remaining image change was contralateral VA stenosis after VA PAO. The incidence of clinical and radiological events was 0.95 and 1.1% per patient-year, respectively.

**Conclusions:** Once the patients surpass the acute phase after PAO, their mid-to-long term course was stable. The risk of late stroke or de novo aneurysm formation was lower than expected in the literature, and the direct comparison to novel reconstructive techniques is warranted in future studies.

**Registration:** https://www.umin.ac.jp/ctr/index.html, trial ID: UMIN000045160

## Introduction

Endovascular parent artery occlusion (PAO) of large intracranial arteries, such as the internal carotid artery (ICA) or vertebral artery (VA), is a well-established treatment option for cerebrovascular diseases, such as dissecting aneurysms, large and giant aneurysms, and vascular injuries. ^1-4^ In these patients, cross-flow via anterior and posterior communicating arteries often plays an important role in perfusing regions previously fed by the occluded artery. To assess the tolerance to PAO, a balloon occlusion test of the corresponding artery is often performed in elective cases.^5^ When the test does not guarantee the tolerance to PAO, bypass surgery can be an option in addition to the PAO.^6,7^

Although PAO is recognized as a standard treatment option, the long-term stability of cerebral blood flow remains a concern. The literature reports the risk of long-term cerebral infarction^3^ and de novo aneurysm formation^8^ during the long-term follow-up after PAO. However, studies focusing on the incidence and outcome of these adverse events with adequate follow-up periods are still insufficient in the literature.

Currently, it has become technically feasible to apply reconstructive endovascular treatment using neck bridging stents and flow diverters to the vascular regions in which endovascular PAO, with or without bypass, was the only treatment option.^9-12^ However, objective insights to choose those novel techniques or conventional PAO for each patient are still lacking. The goal of the present study is to clarify the long-term clinical and radiological outcome of patients who underwent PAO.

Understanding the long-term stability of cerebral blood flow after PAO can help to develop optimal treatment strategies that ensure the best outcomes for patients.

### Patients and methods

This study was designed as a multicenter retrospective observational study. The institutional review board of each participating center approved the study protocol. (Approval number of the primary center: 2021066NI) This work was conducted in accordance with the principle of the Declaration of Helsinki. The requirement of written informed consent was waived because of the study’s retrospective nature. Patients were given the opportunity to opt-out from our hospital website. The study was registered at a Japanese clinical trial database (https://www.umin.ac.jp/ctr/index.html, trial ID: UMIN000045160).

This study included patients who received PAO of their ICA or VA for any purpose between April 2011 and March 2022. We reviewed patient characteristics including age, sex, original pathology requiring PAO, comorbidity, preoperative medications, and preoperative modified Rankin scale (mRS). Details of the endovascular treatment, including preoperative balloon occlusion test assessment and concomitant surgical procedures, such as ventricular drainage and bypass, were reviewed. Any stroke or transient ischemic attack (TIA) within 30 days after PAO was defined as early stroke, and the occurrence rate was analyzed in the ICA PAO group and VA PAO group, respectively.

The procedural details of PAO were up to each participating center. In general, the occlusion of all the affecting regions using detachable coils was intended. For example, if the target pathology was the dissecting aneurysm, occlusion of all the regions covering the aneurysm was performed from the distal and to the proximal normal artery. The use of antithrombotic agents was up to each participating center on case-by-case basis. Basically, heparin was administered systemically during the guiding catheter navigation to maintain the activated clotting time 1.5-2 times longer than its normal value. For elective treatments, at least one oral antiplatelet drug was administered in the perioperative period to prevent potential ischemic complications. In balloon occlusion tests, the target artery was occluded with a balloon catheter for 20 minutes to assess the clinical tolerance to permanent occlusion. Stump pressure from the tip of the balloon catheter and the venous phase delay were also assessed as a reference information to measure the collateral flow more objectively, but the final decision whether to perform PAO was made by each center on a case-by-case basis. When the PAO was considered not feasible according to the results of the balloon occlusion test, or when normal branches arose from the planned occlusion sites, surgical trapping of the region with bypass surgery was considered.

The follow-up information was also analyzed. Patients who did not have a follow-up period of more than 12 months were excluded from this analysis. The clinical follow-up period length, any stroke, TIA, or death during the follow-up period, and the mRS at the last follow-up were retrospectively reviewed from medical records of each participating center. If the patient had vascular imaging tests (computed tomography angiography, magnetic resonance angiography, or catheter cerebral angiography) during their follow-up, those tests were also reviewed at each center to check if new vascular regions like occluded vessel recanalization, intracranial artery stenosis, occlusion, and de novo aneurysm formation developed.

We drew Kaplan-Meyer curves for both clinical and radiological follow-up to analyze the middle-to-long term effect of PAO, using R version 4.1.1 (The R Foundation for Statistical Computing, Vienna, Austria; http://www.R-project.org/). Continuous variables are expressed as mean ± standard deviation, and categorical data are expressed as the number of subjects (percentage of the total).

## Results

A total of 104 cases from eight centers were included in this study. The basic patient characteristics are summarized in **Table1**. The average age of the patients was 52.9±12.6 years old, and males comprised 73 (70.2%) cases. 95 (91.3%) cases were therapeutic VA occlusion. The most frequent pathology necessitating VA PAO was ruptured VA dissection in 73 cases, followed by unruptured VA aneurysm (17 cases), trauma (four cases), and tumor (one case). The remaining nine cases were ICA occlusion. The reason for ICA PAO was unruptured intracranial aneurysm in two cases, ruptured aneurysm in one case, carotid-cavernous fistula in one case, brain tumor in two cases, and iatrogenic ICA injury in three cases. Regarding comorbidity, 42 (41.1%) patients had hypertension, and 13 (12.5%) patients took antiplatelet drugs.

**Table1:** Summary of patient characteristics. DM: diabetes mellitus, HT: hypertension, ICA: internal carotid artery, IHD: ischemic heart disease, VA vertebral artery

|  |  |
| --- | --- |
| **N** | **104** |
| Age | 52.9±12.6 |
| Male | 73 (70.2%) |
| **Occluded artery** |  |
| ICA | 9 (8.7%) |
| VA | 95 (91.3%) |
| **Comorbidity** |  |
| HT | 42 |
| DM | 2 |
| Stroke | 2 |
| IHD | 2 |
| **Preoperative medication** |  |
| Antiplatelet | 13 |
| Anticoagulant | 1 |

Regarding the details of the treatment, preoperative ischemic tolerance was checked with a balloon occlusion test in 15 (14.4%) cases (six cases of ICA PAO and nine cases of VA PAO). No cases underwent concomitant bypass surgery in this cohort. The PAO treatment was successful in all cases. The early (within 30 days) stroke or TIA occurred in 34 (32.7%) cases (32 cases in VA PAO and two cases in ICA PAO).

The ischemic stroke (30 cases) comprised the largest group of early stroke. The remaining four cases were TIA, intraoperative target aneurysm rupture, delayed target aneurysm rupture two weeks after PAO, and intracerebral hemorrhage related to the concomitant ventricular drainage surgery, respectively. As for PAO of the unruptured intracranial aneurysms (N=19, 17 cases in VA PAO and two cases in ICA PAO), the ischemic stroke occurred in 3 cases (16%). No hemorrhagic stroke or TIA was experienced in PAO of unruptured intracranial aneurysms. The details of the ICA and VA PAO are summarized in **Table 2**.

**Table 2:** Details and early outcome of PAO.

| **Occluded artery** | **VA** | **ICA** | **Total** |
| --- | --- | --- | --- |
| **N** | **95** | **9** | **104** |
| **Original Pathology** |  |  |  |
| Ruptured aneurysm | 73 (77%) | 1 (11%) | 74 (71%) |
| UIA | 17 (18%) | 2 (11%) | 19 (18%) |
| Trauma | 4 (4.2%) | 0 (0%) | 4 (3.8%) |
| Tumor | 1 (1.1%) | 2 (22%) | 3 (2.9%) |
| Iatrogenic | 0 (0%) | 3 (33%) | 3 (2.9%) |
| CCF | 0 (0%) | 1 (11%) | 1 (1.0%) |
| **Preoperative BOT** | **9 (9.5%)** | **6 (67%)** | **15 (14%)** |
| **Concomitant bypass surgery** | 0 (0%) | 0 (0%) | 0 (0%) |
| **Stroke within 30 days** | 31 (33%) | 2 (22%) | 33 (32%) |
| Cerebral infarction | 27 (28%) | 2 (22%) | 29 (28%) |
| Intraoperative rupture | 1 (1.1%) | 0 (0%) | 1 (1.0%) |
| Target aneurysm re-rupture | 1 (1.1%) | 0 (0%) | 1 (1.0%) |
| ICH related to ventricular drainage | 1 (1.1%) | 0 (0%) | 1 (1.0%) |
| **TIA within 30 days** | 1 (1.1%) | 0 (0%) | 1 (1.0%) |

BOT: balloon occlusion test, CCF: carotid-cavernous fistula, ICH: intracranial hemorrhage, PAO: parent artery occlusion, TIA: transient ischemic attack, UIA: unruptured intracranial aneurysm, VA vertebral artery

The clinical follow-up over 12 months was available in 78 cases. During a 49.5 ± 24.3 months follow-up period, two cases in VA PAO group experienced stroke or TIA. One was a TIA 53 months after PAO that did not correspond to posterior circulation insufficiency (further details unavailable), and the other was cerebral infarction that occurred at the contralateral VA region five months after PAO (more details described below). Both events did not lead to the patients’ mRS worsening. The remaining event in this cohort was accidental death not related to the PAO procedure 19 months after PAO. The incidence of the composite clinical outcome (Stroke, TIA, and death) was 0.95% per patient-year. The imaging follow-up was performed in 73 cases. Recanalization of the occluded VA was observed in two cases, one month and nine months after their treatment each. The two recurrent cases were treated endovascularly without symptomatic complications. The remaining imaging change was contralateral atherosclerotic VA stenosis 5 months after VA PAO, which caused cerebellar infarction described above. De novo aneurysm formation was not observed. The incidence of imaging change during follow-up was 1.1% per patient-year. In the ICA PAO group, no delayed stroke, TIA, death, or imaging change during their follow-up was experienced. The Kaplan-Meyer curves of clinical and radiological event-free survival are shown in **Figure 1A** and **1B**. Although no event was observed in the ICA PAO group, there was no statistically significant difference between the ICA PAO group and VA PAO group both for clinical and radiological event-free survival.

**Figure1:**
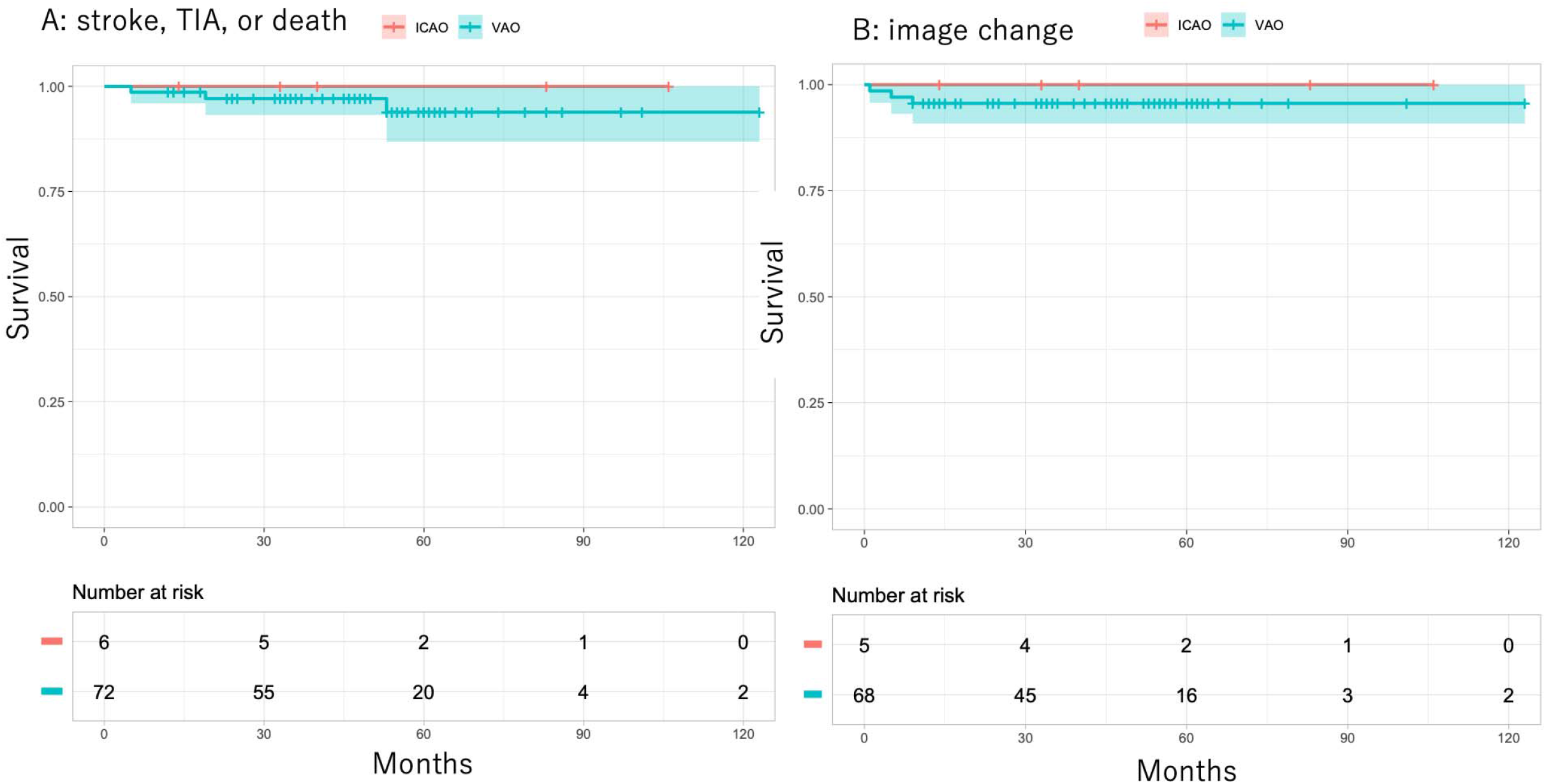
Kaplan-Meyer curve of the clinical (A) and radiological (B) event-free survival during follow-up after parent artery occlusion. Translucent range indicates the 95% confidence interval. ICAO: internal carotid artery occlusion, VAO: vertebral artery occlusion.

## Discussion

In this study, we performed a retrospective assessment of patients’ outcome after PAO of their ICA or VA. PAO was successful in all the cases collected. Once the patients surpassed the early phase after PAO,their mid-to-long term course was stable. Although endovascular PAO is a well-established procedure, its long-term effect has not been well reported in the literature. To the best of our knowledge, our study is one of the largest ones to analyze the mid-to-long term outcome after endovascular PAO of the large intracranial artery, both clinically and radiologically.

As mentioned above, the risk of early stroke (33%) was not negligible in our cohort, and the ischemic stroke was the largest one. One of the reasons for this high complication rate is that most PAOs were performed in an emergency condition like ruptured aneurysm, severe head trauma, or iatrogenic arterial injury. As a result, tolerance to permanent occlusion was checked by a balloon occlusion test in only 15 (14.4%) cases. Proper antithrombotic therapy could sometimes be insufficient in such conditions. In our cohort, although the difference was not statistically significant, the risk of early stroke in the elective unruptured aneurysm PAO (15.7%) was lower. In performing PAO in elective cases, it should be important to check tolerance to permanent artery occlusion with a balloon occlusion test and perform sufficient antithrombotic medical management to prevent perioperative ischemic events. ^5,13,14^ In this study, we included all the periprocedural cerebrovascular events as early stroke, irrespective of their cause-effect relationship to PAO. However, we must recognize that PAO has a certain risk of cerebral ischemia, especially in an acute phase after the procedure.

Initially, this study was designed to clarify the long-term outcome of both ICA and VA PAO. However, the majority of the cases collected (95/104, 91%) was VA PAO. Both the background and the perioperative outcome were quite different between ICA and VA PAO. When we focus on VA PAO, most cases were performed for SAH due to VA dissecting aneurysms and balloon occlusion test was usually not performed in such emergent settings. This emergent nature of VA PAO could lead to the relatively high complication rate in the perioperative period in VA PAO, as described above. On the other hand, we could not collect many ICA PAO cases. In ICA PAO, compared to VA PAO, more cases were performed under elective conditions like unruptured intracranial aneurysms or brain tumor invading ICA, and balloon occlusion test was performed as a preoperative evaluation more frequently. Although not statistically significant due to the small number of ICA PAO cases, the perioperative risk of ICA PAO was lower than that of VA PAO.

In contrast, regarding mid-to-long term follow-up, both ICA and VA PAO showed similar satisfactory stability, both clinically and radiologically. There was only one minor stroke with contralateral VA stenosis during the follow-up period, and this did not lead to mRS worsening. The occluded artery recanalization occurred in two cases within one year after their PAO (one and nine months, respectively), but they were treated with 2^nd^ endovascular treatment without complication. In the literature, as mentioned above, the risk of de novo aneurysm formation after PAO is often referred. The hemodynamic stress to the remaining artery may increase the risk of new aneurysm formation after PAO ^15^. However, we did not encounter de novo aneurysm. To the best of our knowledge, the incidence of de novo aneurysm formation after PAO has not been addressed in large observational studies. The reported risk is largely based on case reports and case series, and some of those studies were performed when noninvasive screening using magnetic resonance imaging was not widespread.^8^ There might be some publication bias in those studies. In addition, in a retrospective cohort study, Tsutsumi et al. exhibited that the annual incidence of de novo aneurysm formation after aneurysm neck clipping was 0.89% in a catheter angiography-based surveillance of 112 patients.^16^ Although the follow-up period was longer in Tsutsumi’s report than the current study (median follow up period 9 versus 4 years), it appears somewhat pessimistic to speculate that the risk of de novo aneurysm formation after PAO is really higher than that of neck clipping of saccular aneurysms. The risk of imaging change after PAO may be too exaggerated in the literature, and those tendency may influence the treatment selection to favor reconstructive endovascular treatment.

Some studies have already compared their result of deconstructive PAO and reconstructive endovascular treatment like stent-assisted coiling and flow diverter stenting. Some emphasize the safety and effectiveness of reconstructive treatment.^12^ In contrast, regarding their mid-to-long term stability, the superiority of endovascular PAO is implied. ^11^ In a large observational study comparing stent-assist coiling and coiling of saccular aneurysms without stents, periprocedural complication rate and re-treatment rate of stent-assisted coiling were 9.4% and 15.5%, respectively.^17^ In a meta-analysis pooling the studies focusing on the outcome of 1186 flow diverter stenting over one year, the delayed ischemic stroke occurred only in one case and the aneurysm recurrence rate was 5%.^18^ However, Studies reporting the feasibility and safety of flow diverters beyond five years are very scarce in the literature. The current study highlights the mid-to-long term effectiveness of therapeutic PAO of intracranial arteries and will serve as an important reference in assessing the safety and effectiveness of stent-assisted coiling, flow diverter, and other novel techniques. Even with the advancement of endovascular treatment devices and techniques, PAO will serve as a considerable therapeutic option when reconstructive technique is not feasible. The direct comparison between PAO and novel reconstructive technique is warranted in the future studies.

There are several limitations to this study. First, the number of participants and the follow-up period are still not sufficient to assess the long-term outcome after PAO. Considering the patients’ age at the treatment, longer follow-up is warranted. However, it is often difficult to follow patients over 10 years in usual clinical practice. Second, the imaging follow-up protocol is not uniform among the participating hospitals, and the assessment of vessel morphology change is performed at each center. Third, the study is basically a single-arm observation, and the comparison between stent-assisted coiling and flow diverter placement is not performed. In addition, we excluded all the patients undertaking direct surgery in the participating centers. The comparison of endovascular PAO and direct surgery with or without bypass should also be warranted. Fifth, information about collateral cerebral blood flow like contralateral VA development and manual compression test of the affecting carotid artery is not collected in this study. We only focused on the preoperative balloon occlusion test, but those simple assessments can also be useful to estimate the blood flow after PAO at least to some extent.

However, despite those limitations, our study highlighted that PAO is a simple procedure and has mid-to-long term stability both clinically and radiologically. In the future, studies comparing our results to the long-term outcome of reconstructive endovascular treatment are awaited to offer the optimal treatment strategy for possible candidates of PAO.

In conclusion, the mid-to-long term stability of patients undergoing PAO of their ICA or VA was exhibited in this multicenter observational study. Although there is a certain risk of early stroke, the patients’ mid-to-long term course was stable. The risk of late stroke or de novo aneurysm formation was lower than expected in the literature. Understanding the long-term stability of cerebral blood flow after PAO can help to offer optimal treatment strategies that ensure the best outcomes for patients. Future studies are warranted to directly compare PAO with reconstructive techniques like stent-assisted coiling and flow diverter stent.

## Data Availability

All the anonymized clinical information of the participants can be offered on request.

## Non-standard Abbreviations and Acronyms

ICA: internal carotid artery
mRS: modified Rankin scale
PAO: parent artery occlusion
TIA: transient ischemic attack
VA: vertebral artery

## Acknowledgements

**none**

